# Using newspapers obituaries to nowcast daily mortality: evidence from the Italian COVID-19 hot-spots

**DOI:** 10.1101/2020.05.31.20117168

**Authors:** Paolo Buonanno, Marcello Puca

**Author notes:** The authors contributed equally to this work. Email addresses:* (Paolo Buonanno), (Marcello Puca).

## Abstract

Real-time tracking of infectious disease outbreaks helps policymakers to make timely data-driven decisions. Official mortality data, whenever available, may be incomplete and published with a substantial delay. We report the results of using newspapers obituaries to nowcast the mortality levels observed in Italy during the COVID-19 outbreak between February 24, 2020 and April 15, 2020. We find that the mortality levels predicted using newspapers obituaries outperforms forecasts based on past mortality according to several performance metrics, making obituaries a potentially powerful alternative source of information to deal with real-time tracking of infectious disease outbreaks.

## 1. Introduction

Since the first suspected pneumonia cases observed on December 2019 in Wuhan (China), the novel coronavirus (COVID-19) causing a severe acute respiratory syndrome turned into a global pandemic.^1^ Having a timely reaction to control the outbreak of an infectious disease is a fundamental factor for the success of a containment measure [1, 2, 3]. While the number of reported cases and infections suffers from several measurement biases, comparing the total mortality rates to those of previous years offers a reliable information on the severity of an epidemic [4, 5]. Mortality data in the middle of a pandemic, however, are not perfect and difficult to estimate [6, 7].^2^ Mortality records, moreover, are published with substantial delay. For example, Britain’s National Statistical Office has recently started to release weekly mortality data after death certificates have been processed.^3^ In Italy, the National Statistical Institute released official mortality data about the January 1, 2020 to February 21, 2020 period only on March 31, 2020, and it usually releases mortality data with a one year lag.^4^

In this paper we propose to use newspapers obituaries as an alternative source of information to ‘now-cast’ daily mortality levels. Specifically, we use obituaries published on the local newspapers of Bergamo and Brescia municipalities, both in the region of Lombardy (Italy), during the Italian COVID-19 outbreak peak, that is from February 24 to May 14, 2020. The Italian region of Lombardy is considered the European hot-spot, with 88,183 reported cases and 15,974 deaths as of May 25, 2020, over a total population of approximately 10 million inhabitants[8, 9].^5^ Figure 1 displays the daily evolution of the raw mortality level (solid line) and the number of published obituaries (dashed line). While obituaries represent only a subset of the officially registered deaths, with a gap increasing at the peak of the outbreak, the correlation between the two measures is glaring.

**Figure 1:**
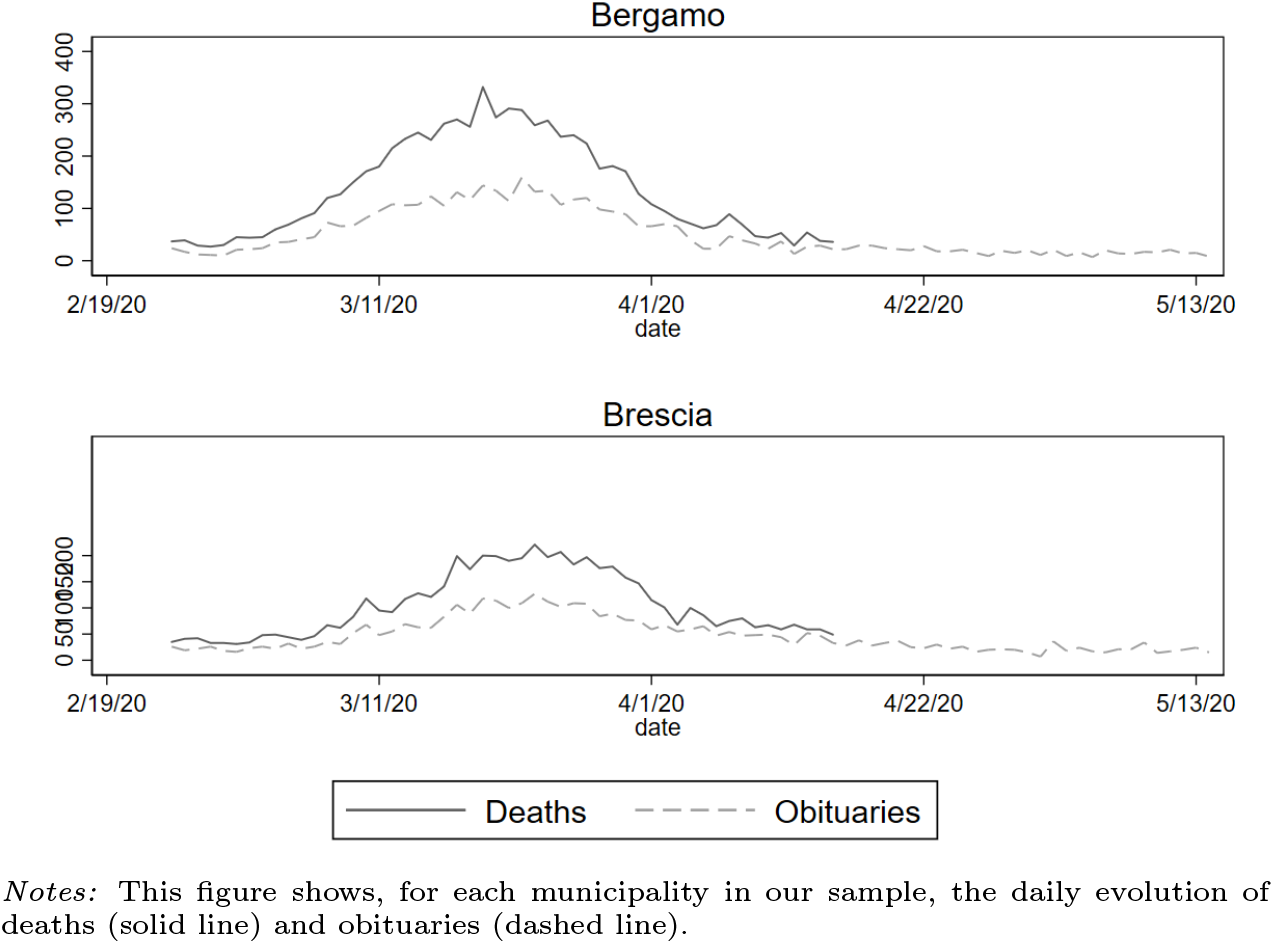
Deaths vs Obituaries

*Our contribution*. Building on standard forecasting techniques, we show the predictive power of newspapers obituaries as an alternative measure of mortality levels. We also compare different forecasting models and report that obituaries-based forecasts outperform all other considered models according to several accuracy criteria.

## 2. Results

Table 1 reports retrospective estimates of daily mortality from February 24, 2020 to May 15, 2020, using several forecasting models, with *Panel A* (resp. *Panel B*) reporting observations for the municipality of Bergamo (resp. Brescia). We compare the estimated mortality level to the true mortality published by ISTAT on May 4, 2020 and computed different accuracy metrics described in 3. These measure include the root mean squared error (RMSE), mean absolute error (MAE), mean absolute percentage error (MAPE), the Theil’s U, the Akaike’s information criterion (AIC), and the Bayesian Information Criterion (BIC). We compare these measures for (i) ordinary least squares (OLS) estimates; (ii) “augmented” autoregressive-moving-average (AARMA(1,2)) estimates with obituaries as exogenous variables; (iii) one lag autoregressive estimates (AR(1)); three lags autoregressive estimates (AR(2)). Comparing these metrics, we report that the AARMA(1,2) model outperforms all other models according to every performance metric, for both municipalities in our sample.

**Table 1:**
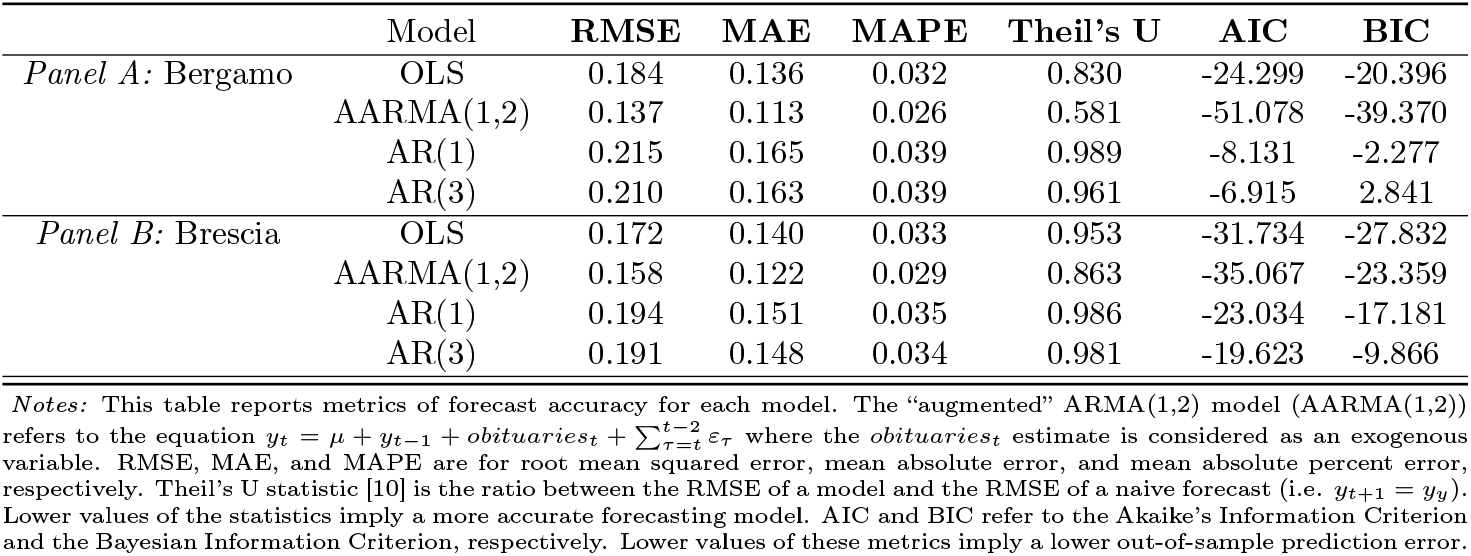
Comparison of different forecasting models of mortality.

|  | Model | RMSE | MAE | MAPE | Theil's U | AIC | BIC |
| --- | --- | --- | --- | --- | --- | --- | --- |
| *Panel A: Bergamo* | OLS | 0.184 | 0.136 | 0.032 | 0.830 | -24.299 | -20.396 |
|  | AARMA(1,2) | 0.137 | 0.113 | 0.026 | 0.581 | -51.078 | -39.370 |
|  | AR(1) | 0.215 | 0.165 | 0.039 | 0.989 | -8.131 | -2.277 |
|  | AR(3) | 0.210 | 0.163 | 0.039 | 0.961 | -6.915 | 2.841 |
| *Panel B: Brescia* | OLS | 0.172 | 0.140 | 0.033 | 0.953 | -31.734 | -27.832 |
|  | AARMA(1,2) | 0.158 | 0.122 | 0.029 | 0.863 | -35.067 | -23.359 |
|  | AR(1) | 0.194 | 0.151 | 0.035 | 0.986 | -23.034 | -17.181 |
|  | AR(3) | 0.191 | 0.148 | 0.034 | 0.981 | -19.623 | -9.866 |

*Notes:* This table reports metrics of forecast accuracy for each model. The “augmented” ARMA(1,2) model (AARMA(1,2)) refers to the equation $y_t = \mu + y_{t-1} + obituaries_t + \sum_{r=1}^{t-2} \varepsilon_r$ where the *obituaries<sub>t</sub>* estimate is considered as an exogenous variable. RMSE, MAE, and MAPE are for root mean squared error, mean absolute error, and mean absolute percent error, respectively. Theil's U statistic [10] is the ratio between the RMSE of a model and the RMSE of a naive forecast (i.e. $y_{t+1} = y_t$ ). Lower values of the statistics imply a more accurate forecasting model. AIC and BIC refer to the Akaike's Information Criterion and the Bayesian Information Criterion, respectively. Lower values of these metrics imply a lower out-of-sample prediction error.

Figure 2 displays the forecasted mortality against the observed mortality level. A close inspection of the estimates shows that both the AARMA(1,2) and the OLS estimates outperform models based only on previously observed mortality data (i.e. AR(1) and AR(3)) over the entire period in our sample. Figure 3 displays the daily evolution of the estimated standard errors for each model. Also in this case, the OLS estimate outperforms the other models during the entire period in our sample.

**Figure 2:**
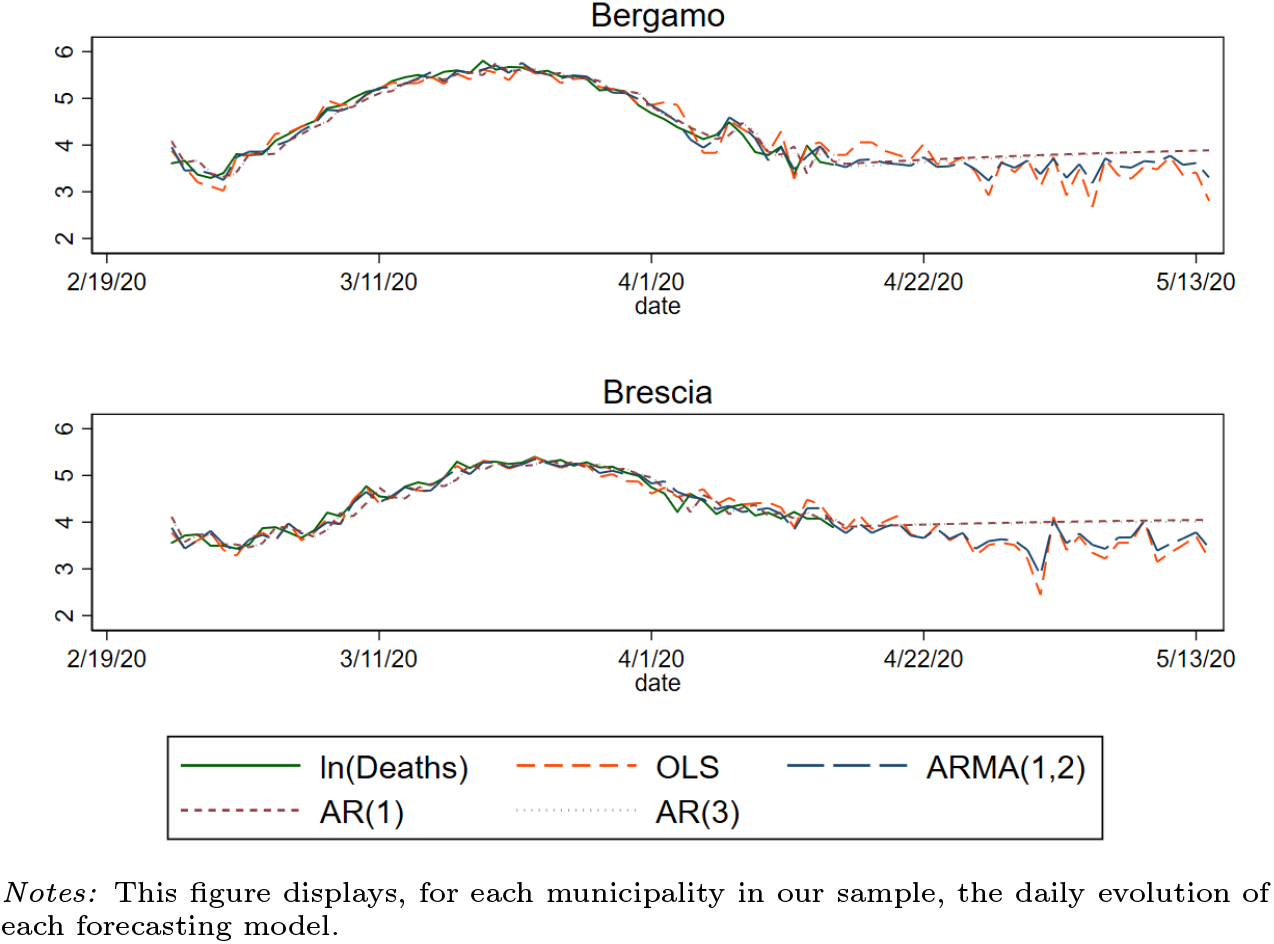
Forecasts

**Figure 3:**
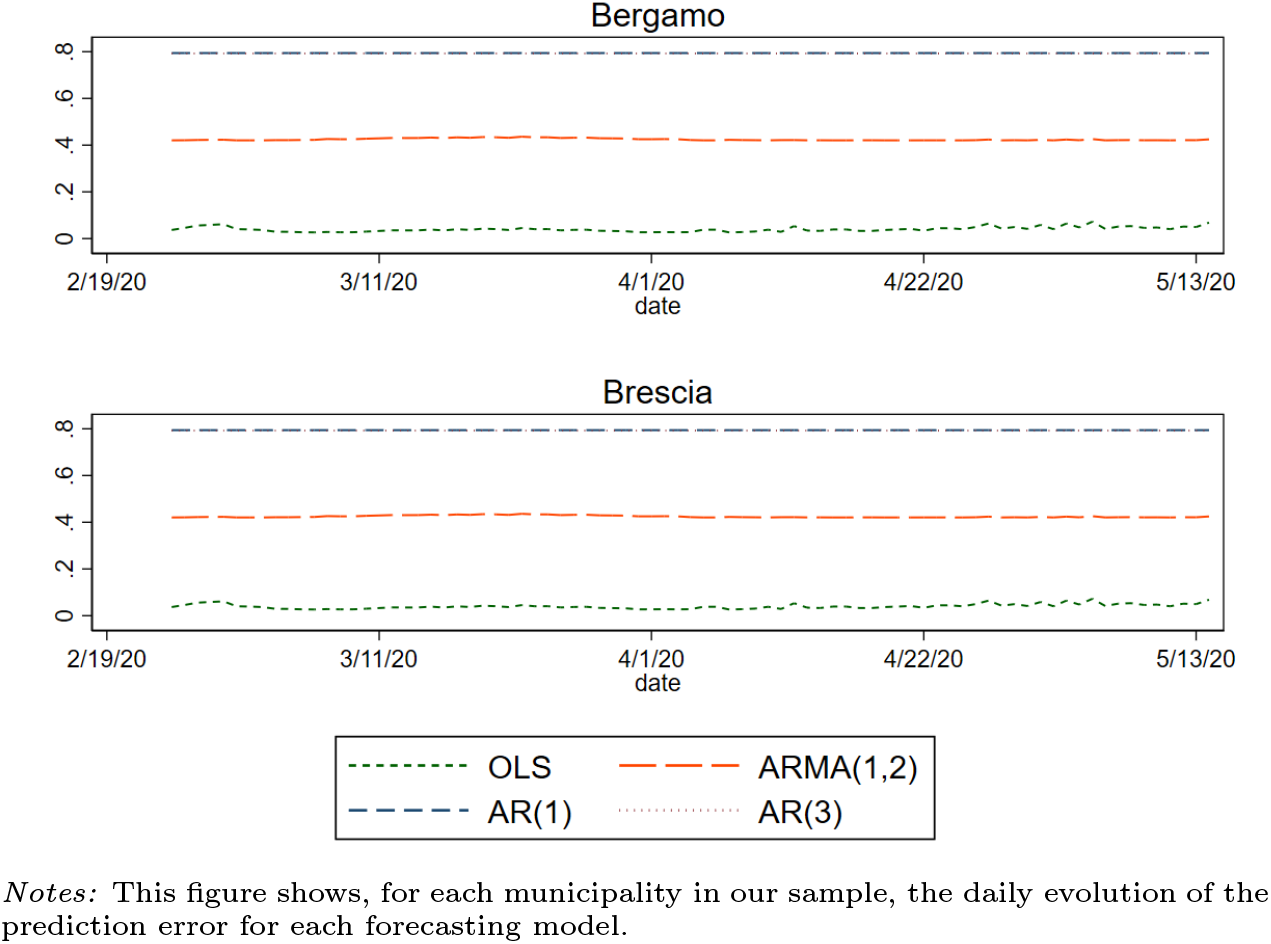
Deaths vs Obituaries

## 3. Data and Methods

The basic principle of now-casting is exploiting information which is published at a higher frequency than the variable of interest [11]. We explore the accuracy of newspapers obituaries published in local newspapers in predicting actual daily mortality in almost real time. Newspapers obituaries contain information on individual characteristics such as name, surname, gender, age, date of death, and the municipality of death. This information allows us to increase the information set available to external observers and estimate a real-time mortality rate.

*Newspapers obituaries*. We digitalized newspapers obituaries published by *L’Eco di Bergamo* and *Il Giornale di Brescia*, the two most read and circulated newspapers in the province of Bergamo and in the province of Brescia, respectively.^6^ Our final dataset contains 4,054 unique individuals from February 24 to May 14, 2020 for the province of Bergamo and 3,784 unique individuals for the province of Brescia over the same period.

We combine obituaries data with mortality data at the municipality level released by the Italian National Statistical Institute (ISTAT) on May 9, 2020.^7^ The ISTAT dataset contains daily deaths at the municipality level from January 1 to April 15, 2020 for a sample of 4,433 Italian municipalities. The ISTAT sample covers the universe of municipalities belonging to the two provinces of our analysis (243 municipalities in the province of Bergamo and 205 municipalities in the province of Brescia).

*Formulation of the AARMA(1,2) model*. Our AARMA(1,2) model is motivated by the inspection of the autocorrelation and partial autocorrelation plots, which display a one lag significant autocorrelation coefficient, and a two lags partial autocorrelation coefficients. This leads us to estimate the following model

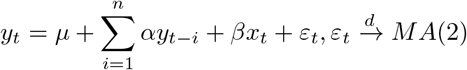

where *y_t_* = ln(*mortality_t_*) is the log-transformed mortality observed at time *t, x_t_* = ln(*obituaries_t_*) is the log-transformed number of newspapers obituaries published at time *t*, which is assumed to be exogenous with respect to the time series {*y_t_*} (i.e. ℰ[*ε_t_*|*x_t_*] = 0).

*Accuracy metrics*. The RMSE, MAE, MAPE, and Theil’s U of the estimator 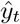 to the target mortality level *y_t_* are defined, respectively, as 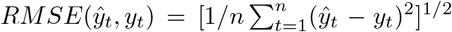, 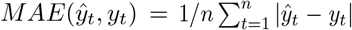, 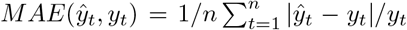, 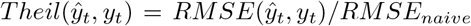, where *RMSE_naive_* refers to the RMSE of a naive forecast, i.e. *y_t_* = *y_t−1_*. The AIC and BIC are defined, respectively, as 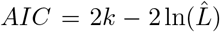 and 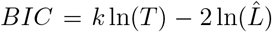, where 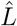 maximizes the likelihood function of the estimated model, *k* is the number of estimated parameters, and *T* is the sample size.

## 4. Discussion and concluding remarks

We use newspapers obituaries to nowcast the mortality levels observed in Italy during the COVID-19 outbreak peak. We find that forecasting models using newspapers obituaries outperform other models based on previously observed mortality. Our approach, despite powerful, is not free from limitations. First, newspapers obituaries may underrepresent the actual mortality level, an issue that becomes more severe during the epidemic peak (see Figure 1). Such underrepresentation, however, goes against our estimates since it should decrease the precision of our estimates. Second, despite concentrated in the most affected Italian region, our sample refers only to two municipalities. We are agnostic about the existence of heterogeneous individual behavioral attitudes towards publishing newspapers obituaries in other locations.^8^ Understanding how such heterogeneity may affect our estimates constitutes a valuable path for future research.

## Data Availability

The data used in this paper are authors' elaboration of publicly available data.

## Acknowledgements

We thank Nunzia Vallini (Director of *Il Giornale di Brescia*) and Mauro Torri (CEO of *Editoriale Bresciana*) for their help. We thank Sergio Galletta for useful comments and discussions. We also thank Endri Avduli and Oumar Ben Salha for research assistance.

## Footnotes

1 World Health Organization rolling updates available at https://www.who.int/emergencies/diseases/novel-coronavirus-2019.

2 There is substantial evidence that the reported number of deaths underestimates the actual mortality value, c.f. https://www.nytimes.com/interactive/2020/04/21/world/coronavirus-missing-deaths.html, https://www.nationalgeographic.com/science/2020/05/what-we-need-to-find-true-coronavirus-death-toll/.

3 C.f. https://www.ons.gov.uk/peoplepopulationandcommunity/birthsdeathsandmarriages/deaths/bulletins/deathsregisteredweeklyinenglandandwalesprovisional/weekending20march2020.

4 See Section 3 for further details.

5 Data on cumulative cases are available at http://www.protezionecivile.gov.it/media-communication/press-release/detail/-/asset_publisher/default/content/coronavirus-la-situazione-dei-contagi-in-ita-37.

6 In 2019, the daily number of readers of *L’Eco di Bergamo* has been 402,000, while the daily number of readers of *Il Giornale di Brescia* has been 427,000. Source: http://audipress.it/quotidiani/

7 Data are available at the ISTAT website: https://www.istat.it/it/archivio/240401.

8 The large heterogeneity observed in civic attitude and prosocial behavior across Italian municipalities may play a role in determining such propensity to publish obituaries [12].

